# Interobserver Agreement of Lung Ultrasound Findings of COVID-19

**DOI:** 10.1101/2020.08.16.20176156

**Authors:** Andre Kumar, Yingjie Weng, Sally Graglia, Sukyung Chung, Youyou Duanmu, Farhan Lalani, Kavita Gandhi, Viveta Lobo, Trevor Jensen, Jeffrey Nahn, John Kugler

## Abstract

**Background:** Lung ultrasound (LUS) may be used in the diagnostic evaluation of patients with COVID-19. An abnormal LUS is associated with increased risk for ICU admission in COVID-19. Previously described LUS manifestations for COVID-19 include B-lines, consolidations, and pleural thickening. The interrater reliability (IRR) of these findings for COVID-19 is unknown.

**Research Question:** What is the interrater reliability of lung ultrasound findings in patients with RT-PCR confirmed COVID-19?

**Study Design and Methods:** This study was conducted at conducted at two academic medical centers between 03/2020-06/2020. Nine physicians (hospitalists: n=4; emergency medicine: n=5) independently evaluated n=20 LUS scans (n=180 independent observations) collected from RTPCR confirmed COVID-19 patients. These studies were randomly selected from an image database consisting of COVID-19 patients evaluated in the emergency department with portable ultrasound devices. Physicians were blinded to any patient information or previous LUS interpretation. Kappa values (κ) were used to calculate IRR.

**Results:** There was substantial IRR on the following items: normal LUS scan (κ=0.79 [95% CI: 0.72-0.87]), presence of B-lines (κ=0.79 [95% CI: 0.72-0.87]), >=3 B-lines observed (κ=0.72 [95% CI: 0.64-0.79]). Moderate IRR was observed for the presence of any consolidation (κ=0.57 [95% CI: 0.50-0.64]), subpleural consolidation (κ=0.49 [95% CI: 0.42-0.56]), and presence of effusion (κ=0.49 [95% CI: 0.41-0.56]). Fair IRR was observed for pleural thickening (κ=0.23 [95% CI: 0.15-0.30]).

**Interpretation:** Many LUS manifestations for COVID-19 appear to have moderate to substantial IRR across providers from multiple specialties utilizing differing portable devices. The most reliable LUS findings with COVID-19 may include the presence/count of B-lines or determining if a scan is normal. Clinical protocols for LUS with COVID-19 may require additional observers for the confirmation of less reliable findings such as consolidations.

**Clinicaltrials.gov Registration:** NCT04384055

**Disclosures:** Andre Kumar, MD, MEd is a paid consultant for Vave Health, which manufactures one of the ultrasound devices used in this study. His consultant duties include providing feedback on product development. The other authors do not have any items to disclose.

## Introduction

Point-of-care ultrasound (POCUS) has the potential to transform healthcare delivery in the era of COVID-19 with its diagnostic expediency.^1^ POCUS provides real-time interpretation of a patient’s condition in an augmented clinical manner, which can immediately impact management decisions.^2,3^ POCUS devices, particularly handheld devices, are often cheaper than traditional radiological equipment such as X-ray or computerized tomography (CT) machines, which makes POCUS ideal for COVID-19 surge scenarios where these resources may be limited.^4,5^ Furthermore, POCUS may reduce the number of providers exposed to patients with COVID-19 by decreasing the need for radiological studies, which could result in decreases of personal protective equipment usage by radiological technicians and the resources needed to decontaminate larger radiological equipment.^4^

Previously reported pulmonary manifestations of COVID-19 with POCUS include B-lines, subpleural consolidations, pleural thickening, and absence of pleural effusions.^4–8^ POCUS has been proposed to aid in the diagnosis of COVID-19,^9^ as well as to predict intensive care unit (ICU) admission or death.^10^ Given the potential for POCUS to predict outcomes among COVID-19 patients, there is a significant need to determine if these findings can be reliably interpreted among providers. Outside of COVID-19, previous investigations have found moderate to excellent interobserver agreement for many lung ultrasound (LUS) findings, including B-lines, consolidations, and effusions.^2,11,12^

In this study, we characterize the interobserver agreement of LUS findings that have been described for COVID-19. These images were collected using portable ultrasound devices, which may be more commonly utilized as a result of the COVID-19 pandemic.

## Methods

### Participants & Setting

This analysis was conducted as part of an ongoing study investigating the role of POCUS for COVID-19 that is being conducted at four medical centers in the United States (http://Clinicaltrials.gov Registration: NCT04384055). This investigation utilizes a prospectively collected database and includes adult patients who meet the following criteria: 1) presentation to the emergency department with symptoms^13^ suspicious for COVID-19, 2) a positive nasopharyngeal RT-PCR for SARS-CoV-2, and 3) received a lung ultrasound (LUS) during their emergency department course or subsequent hospitalization (up to 28 days from admission). This study has received institutional review board (IRB) approval at all participating sites.

### Study Procedure

In this study, a total of n=20 LUS scans collected between 3/2020 and 6/2020 were randomly selected from our image database consisting of COVID-19 POCUS images. These scans originated from n=13 patients, which occurred because some patients received multiple scans during a 28-day period. Any scans from the same patient were acquired on different days. All scans were collected using a 12-zone lung protocol, which we have previously described and is also demonstrated in Figure 1.^6^ Each zone clip was 6 seconds in length.^6^ The LUS studies were collected on the following devices: Butterfly iQ (n=9), Vave Personal Ultrasound (n=6), Philips Lumify (n=4), and Sonosite M-turbo (n=1), which represent the commercially-available portable devices at our institutions. In our study protocol, providers can acquire these scans using any portable device available to them, and the types of devices used in this analysis were the result of random selection from our database.

**Figure 1.**
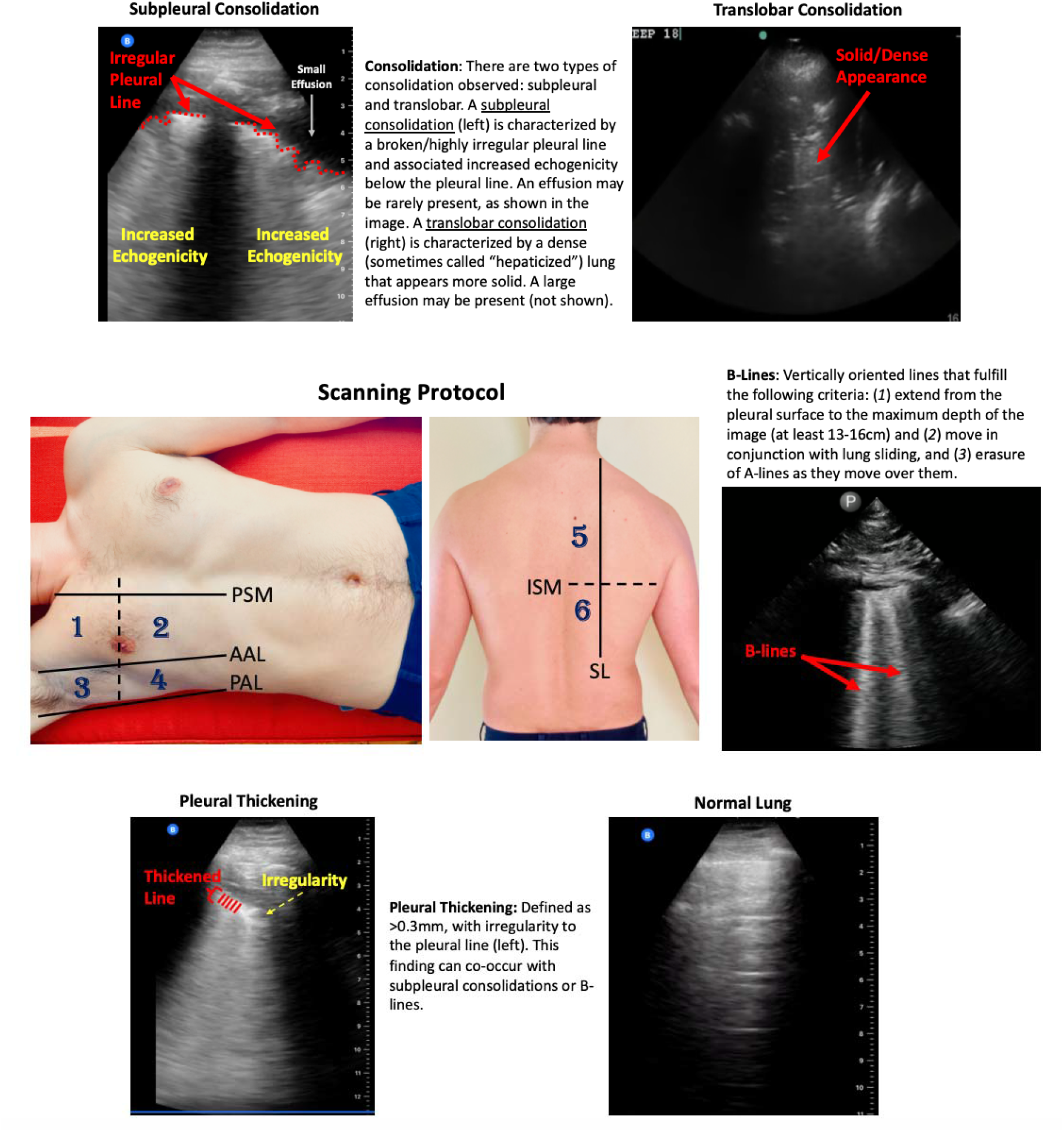
Scanning Protocol and Lung Ultrasound Findings in COVID-19 Patients. This study utilized a 12-zone protocol.^27^ On each hemithorax, there are 6 zones. The exam begins on the patient’s right side. Zones 1-2 (anterior zones) are between the parasternal margin (PSM) and the he st zones) are between the anterior axillary line (AAL) and and are best obtained in the mid-clavicular line. Zones 3-4 (lateral zones) are between the anterior axillary line (AAL) and posterior axillary line (PAL) and are best obtained in the mid-axillary line. The nipple line serves as a bisecting area between these zones. Zones 5–6 (posterior zones) are medial to the scapular line (SL) and are bisected by the inferior scapular margin (ISM). The zone areas are repeated on the contralateral hemithorax (starting with zone 7). This figure contains an overview of the observed ultrasound findings based on previously described terminology.^3,27,28^

This study included nine physicians from two specialties (hospitalists n=4; emergency medicine, n=5) who are regular users of LUS and have leadership positions in POCUS at their respective institutions. These nine physicians independently evaluated n=20 LUS scans (n=180 independent observations). The physicians were blinded to any patient information or previous interpretation. All physicians received a document that contained the scanning protocol (Figure 1) as well as definitions for each of the pathological findings (Figure 1). The physicians met for a 1-hour calibration session at the beginning of the study to review sample videos and discuss their real-time interpretations. These samples were not included in the interrater analysis. The physicians were then instructed to independently assess the 20 de-identified studies and input their interpretation using a central electronic database (REDCap).^14^ No other instruction on image interpretation was provided to the group during the independent assessment period.

### Statistical Analysis

The degree of agreement for Kappa values were based on the research originally described by Cohen and later Landis & Koch.^15–17^ In our study, Fleiss kappa statistics and corresponding 95% confidence intervals were reported. In this scheme, we interpreted the Fleiss’ kappa statistics^17^ with the following criteria: κ< 0 (no agreement), κ=0–0.20 (none to slight agreement), κ=0.21-0.40 (fair agreement), κ=0.41–0.60 (moderate agreement), κ=0.61-0.80 (substantial agreement), κ=0.81–1.0 (near perfect agreement). All analyses were performed using R statistical programming language, version 3.6.1 (Vienna, Austria).

## Results

### A. Study Population

Patient demographics and clinical characteristics are described in Table 1. The mean body mass index (BMI) for the patient population was 30.5 kg/m^2^ (range 24.6-37.5). The mean age for the study cohort was 49.2 (SD 19.2). Approximately 57% of the patients were female (Table 1)

**Table 1.** Patient Characteristics. N=20 scans were randomly selected from our prospectively acquired database, which originated from N=13 patients. The patient characteristics are displayed. BMI, Body Mass Index; ICU, Intensive Care Unit, COPD, Chronic Obstructive Pulmonary Disease.

Tables/Figures
|  |  |
| --- | --- |
| Mean Age (SD) | 49.2 (19.2) |
| Mean BMI (kg/m<sup>2</sup>; SD) | 30.5 (3.9) |
| Female (%) | 7 (54%) |
| Admitted to ICU (%) | 6 (46%) |
| Medical History |  |
| Hypertension | 3 (23%) |
| Hyperlipidemia | 3 (23%) |
| Diabetes | 5 (38%) |
| Coronary Artery Disease | 2 (15%) |
| Heart Failure | 0 (0%) |
| COPD | 0 (0%) |
| Asthma | 3 (23%) |
| Malignancy | 0 (0%) |

### B. Normal Vs. Abnormal Scan

Overall, there was substantial agreement on determining whether a scan contained no abnormalities, including absence of B-lines, consolidations, pleural thickening, or effusions (κ=0.79 [95% CI: 0.72-0.87]; Table 2).

**Table 2.** Interobserver Agreement of Lung Ultrasound Findings Among COVID-19 Patients. The degree of agreement was based on previously reported methodology.^17^ Effusion size was defined as < 1cm from the visceral and parietal pleura vs. >=1 cm.

| Finding | Kappa Value | 95% CI | Degree of Agreement |
| --- | --- | --- | --- |
| Normal Examination | 0.79 | [0.72-0.87] | Substantial Agreement |
| B-lines Present | 0.79 | [0.72-0.87] | Substantial Agreement |
| 3 or More B-Lines per Lung Field | 0.72 | [0.64-0.79] | Substantial Agreement |
| Consolidation (Translobar or Subpleural) | 0.57 | [0.50-0.64] | Moderate Agreement |
| Subpleural Consolidation | 0.49 | [0.42-0.56] | Moderate Agreement |
| Pleural Effusion | 0.49 | [0.41-0.56] | Moderate Agreement |
| Effusion Size (Small Vs. Moderate) | 0.47 | [0.40-0.55] | Moderate Agreement |
| Pleural Thickening | 0.23 | [0.15-0.30] | Fair Agreement |
| Total Count of B-Lines per Scan | 0.16 | [0.14-0.17] | None to Slight Agreement |
| Translobar Consolidation | 0.15 | [0.08-0.23] | None to Slight Agreement |

### C. B-Lines

There was substantial interrater agreement on the presence of B-lines (κ=0.79 [95% CI: 0.72-0.87]) or whether the scan contained three or more B-lines per lung field (κ=0.72 [95% CI: 0.64–0.79]; Table 2). The presence of bilateral B-lines also demonstrated substantial agreement (κ=0.70 [95% CI: 0.63-0.78]; Table 3). Similarly, the presence of B-lines had substantial agreement across the anterior (κ=0.79 [95% CI: 0.72-0.87]), lateral (κ=0.76 [95% CI: 0.69-0.83]), and posterior lung zones (κ=0.70 [95% CI: 0.63-0.77]; Table 3). Notably, the total count of B-lines per scan had slight agreement (κ=0.16 [95% CI: 0.14-0.17]).

### D. Consolidation

There was moderate interrater agreement on the presence of any consolidation (κ=0.57 [95% CI: 0.50-0.64]; Table 2). When analyzed by consolidation subtype, the presence of subpleural consolidations had moderate agreement (κ=0.49 [95% CI: 0.42-0.56]) and translobar consolidations had slight agreement (κ=0.15 [95% CI: 0.08-0.23]). The presence of bilateral consolidation had fair agreement (κ=0.28 [95% CI: 0.20-0.35]; Table 3). There was variable agreement of the presence of any type of consolidation by location: anterior (κ=0.71 [95% CI: 0.63-0.78]), lateral (κ=0.56 [95% CI: 0.48-0.63]), and posterior (κ=0.86 [95% CI: 0.78-0.93]; Table 3).

**Table 3.** Interobserver Agreement of Lung Ultrasound by Lung Field Location. Lung zones (anterior, lateral, and posterior) are described in Figure 1.

| Finding | Kappa Value | 95% CI | Degree of Agreement |
| --- | --- | --- | --- |
| **B-Lines** |  |  |  |
| Anterior B-Lines | 0.79 | [0.72-0.87] | Substantial Agreement |
| Lateral B-Lines | 0.76 | [0.69-0.83] | Substantial Agreement |
| Posterior B-Lines | 0.70 | [0.63-0.77] | Substantial Agreement |
| Bilateral B-lines | 0.70 | [0.63-0.78] | Substantial Agreement |
| **Consolidation** |  |  |  |
| Anterior Consolidation | 0.71 | [0.63-0.78] | Substantial Agreement |
| Lateral Consolidation | 0.56 | [0.48-0.63] | Moderate Agreement |
| Posterior Zone Consolidation | 0.86 | [0.78-0.93] | Near Perfect Agreement |
| Presence of Bilateral Consolidation | 0.28 | [0.20-0.35] | Fair Agreement |
| **Pleural Thickening** |  |  |  |
| Anterior Pleural Thickening | 0.33 | [0.26-0.40] | Fair Agreement |
| Lateral Pleural Thickening | 0.34 | [0.26-0.41] | Fair Agreement |
| Posterior Pleural Thickening | 0.48 | [0.41-0.55] | Moderate Agreement |
| Presence of Bilateral Pleural Thickening | 0.33 | [0.26-0.41] | Fair Agreement |
| **Effusions** |  |  |  |
| Presence of bilateral effusion | 0.40 | [0.32-0.47] | Fair Agreement |

### E. Pleural Thickening

There was fair agreement on the presence of pleural thickening (κ=0.23 [95% CI: 0.15-0.30]; Table 2). Similarly, there was fair agreement on the presence of bilateral pleural thickening (κ=0.33 [95% CI: 0.26-0.41]). When analyzed by location, pleural thickening demonstrated fair to moderate agreement across the anterior, lateral, and posterior zones (Table 3).

### F. Pleural Effusion

There was moderate agreement on the presence of pleural effusions (κ=0.49 [95% CI: 0.41-0.56]) and size of effusion (κ=0.47 [95% CI: 0.40-0.55]; Table 2). The presence of bilateral pleural effusions had fair agreement (κ=0.40 [95% CI: 0.32-0.47]; Table 3).

## Discussion

In this study, we investigated the interobserver agreement of LUS findings that have been previously described with COVID-19. Several LUS findings demonstrated substantial agreement (e.g. B-lines), while others demonstrated moderate to fair agreement (e.g. consolidations, pleural thickening, or effusions). This study represents the first investigation of the interobserver agreement of LUS findings in COVID-19 and includes practitioners from multiple specialties who utilized several portable devices.

There are several implications of these findings. Previous authors have demonstrated the presence of B-lines can be used in the diagnostic evaluation for COVID-19.^6,9^ Others have proposed that bilateral B-lines on LUS increase the risk of ICU admission or death with COVID-19.^10^ Our results suggest that there is substantial interobserver agreement for the presence of B-lines across multiple provider specialties utilizing different handheld devices. Therefore, the presence of B-lines may represent a reliable, diagnostic, and prognostic clinical entity for COVID-19. Similarly, there was substantial agreement on whether a LUS scan was interpreted as abnormal vs. normal. While the prognostic value of a normal LUS for COVID-19 remains uncertain, others have shown that chest radiograph abnormalities with COVID-19 are associated with ICU admission.^18^ Furthermore, an abnormal LUS scan (outside of COVID-19) has prognostic implications across multiple diseases.^19–21^ Future studies are needed to determine if LUS can reliably predict clinical outcomes with COVID-19.

How do these findings compare to the interrater reliability literature for LUS outside of COVID-19? Previous investigations have demonstrated moderate to substantial agreement for B-lines^22–25^. In contrast, there is only moderate to fair agreement for consolidation^11,12,22^, pleural irregularity^^22^^, and effusions.^11,22^ Others have shown that there is substantial interrater reliability for LUS across differing probes (e.g. linear vs. phased array)^25,26^, which is important given different portable devices were used in this investigation. Future investigations of LUS for COVID-19 should consider multiple observers to confirm less-reliable findings and utilize a standardized interpretation protocol. This latter point may be important because there are variable definitions of LUS findings in the literature, especially for consolidations.^5,11,12,22^. Although consolidations had moderate agreement in this study, the reliability of this finding might improve with more specified definitions and consensus-based guidelines.

There are several limitations to this study. Our study population was confined to patients who presented to the emergency department or were hospitalized, which limits the generalizability of these findings for providers in the outpatient setting. Although we randomly selected LUS studies from our database, we sampled fewer patients (n=13) than the total number of scans analyzed (n=20). The researchers in this study completed a 1-hour calibration session and had a definition sheet when interpreting images. Therefore, the IRR may be lower for certain findings among less-trained practitioners. Although this study utilized several portable devices, there was variability in the image quality of these devices (particularly when visualizing the pleural line), which may have affected the results regarding pleural thickening or subpleural consolidation. Despite these limitations, this study represents one of the first dedicated investigations into the interobserver agreement of LUS findings for COVID-19.

In conclusion, many LUS manifestations for COVID-19 appear to have substantial to moderate IRR across providers from multiple specialties utilizing differing portable devices. More reliable findings included the determination of a normal scan, the presence and location of B-lines, and determining if >=3 B-lines were present. Less reliable findings related to the presence or locations of consolidations, pleural thickening, or effusions. Since presence of B-lines may have diagnostic and prognostic utility for COVID-19, this finding can likely be interpreted without additional oversight and can be incorporated into future clinical protocols. In contrast, other findings such as pleural thickening may be less reliable, and clinical protocols incorporating these findings may require quality assurance for accurate interpretation.

## Data Availability

Data is available on request to the corresponding author.

## Acknowledgements

Andre Kumar, MD, MEd, Yingjie Weng, MS, Sally Graglia, MD, MPH, Sukyung Chung, PhD, Youyou Duanmu, MD, MPH, Farhan Lalani, MD, Kavita Gandhi, MD, PhD, Viveta Lobo, MD, Trevor Jensen, MD, Jeffrey Nahn, MD, and John Kugler, MD all contributed equally to study design, data collection, data interpretation, data analysis, and manuscript writing.

